# In vivo assessment of cardiac radiofrequency ablation in a large-animal model using photoacoustic-ultrasound imaging

**DOI:** 10.1101/2024.09.18.24313514

**Authors:** Nilesh Mathuria, Krithik Vishwanath, Blake C. Fallon, Antonio Martino, Giorgio Brero, Richard C. Willson, Miguel Valderrabano, Carly S. Filgueira, Richard R. Bouchard

**Affiliations:** Houston Methodist Heart and Vascular Center, Houston Methodist Research Institute, Houston Methodist Hospital, Houston, Texas, USA; Department of Imaging Physics, The University of Texas MD Anderson Cancer Center, Houston, Texas, USA; Department of Nanomedicine, Houston Methodist Research Institute, Houston, Texas, USA; Department of Materials Science and Engineering, University of Houston, Houston, Texas, USA; Department of Electronics, Politecnico di Torino, Torino, Italy; Department of Biochemical & Biophysical Sciences, University of Houston, Houston, Texas, USA; Department of Cardiovascular Surgery, Houston Methodist Research Institute, Houston, Texas, USA; The University of Texas MD Anderson Cancer Center UTHealth Graduate School of Biomedical Sciences, Houston, Texas, USA

**Author notes:** Equal contribution (Nilesh Mathuria, Krithik Vishwanath). Corresponding Author: Richard R. Bouchard, 1881 East Rd., Houston, TX 77054. Co-corresponding Author: Carly S. Filgueira, 6670 Bertner Ave., Houston, TX 77030.

## Abstract

One of the fundamental unmet clinical needs within cardiac electrophysiology is intraoperative assessment of catheter ablation, which can lead to recurrent arrhythmias and subsequent complications if ineffective. This work demonstrates photoacoustic imaging (PAI) of radiofrequency ablation (RFA) lesions in an *in vivo* swine model (n=3). Spectral unmixing of PAI data provides local myocardial characterization (e.g., oxygen saturation & tissue ablation) by overlaying unmixed PAI images with B-mode ultrasound imaging (PAI/US), with the latter providing anatomical context. Based on stained gross pathology, areas of central tissue necrosis coincided with increases in unmixed ablated regions of the myocardium. An average contrast-to-noise ratio of 2.8±0.2 confirmed lesion detectability, while the lesion dimensions quantified from PAI and pathology did not present significant differences. *In vivo* PAI of RFA lesions to determine ablation characteristics could lead to a paradigm shift in catheter ablation assessment and improve clinical outcomes.

---

Radiofrequency ablation (RFA) remains the most common modality for catheter ablation (CA) of medically refractory cardiac arrhythmias. Success rates remain suboptimal, however, with 30% ventricular tachycardia (VT) recurrence within one year.^1^ Such recurrence is often due to ineffective ablation, which occurs when tissue temporarily loses conduction but later “recovers.” Despite advancements in RFA guidance, accurate intraoperative lesion assessment remains elusive. Therefore, an unmet clinical need exists to provide intraoperative lesion assessment during CA procedures to avoid inadequate or excessive ablation. Photoacoustic imaging (PAI) can address this need by providing local myocardial characterization (e.g., deoxy-hemoglobin [Hb] & oxy-hemoglobin [HbO_2_]) at depth. PAI can also be overlaid with “B-mode” ultrasound imaging (PAI/US) to provide anatomical context. *Ex vivo* investigations using PAI have demonstrated spectral distinctions in Hb (i.e., elimination of ~760-nm local maximum) between ablated (Hb_A_) and non-ablated regions of myocardium.^2^ Consequently, an *in vivo* porcine study was conducted to investigate PAI’s translational potential for intraoperative RFA lesion assessment.

RFA lesions were created on the mid-to-apical, anterior left ventricle of 3 Domestic Swine (female; 39-45 kg; 3-5 mos; Oak Hill Genetics) with an open-chest preparation (Fig. 1A) using a Thermocool STSF™ irrigated catheter (30 W for 30 s; 17 cc/min; 10-20 gm contact force; 90-100 bpm). Animal sedation occurred via intramuscular injection of 20 mg/kg ketamine, 0.5 mg/kg midazolam, & 0.1 mg/kg hydromorphone and intravenous administration of 0.04 mg/kg atropine; infusions of lidocaine (0.5-1 mg/min; arrhythmia suppression) & norepinephrine (1-4 mcg/min; blood-pressure support) were given as needed. Following ablation, PAI/US was performed using a Vevo 2100-LAZR with an LZ-250 linear-array transducer (21-MHz center frequency; 256 elements) using a custom, 3D-printed suction coupler to mitigate transducer motion (green arrow Fig.1A). Matched PAI and US images were acquired (~200 ms) without averaging for each wavelength (740, 755, 760, 780, 800, & 850 nm), which alternated sequentially to complete an ensemble (top Fig.1B) and then repeated for ~1 min. After euthanasia, hearts were excised, sliced along the imaging plane (perpendicular to surface), placed in 1% triphenyl tetrazolium chloride (15 min @ 37°C), then photographed.

**Figure 1.**
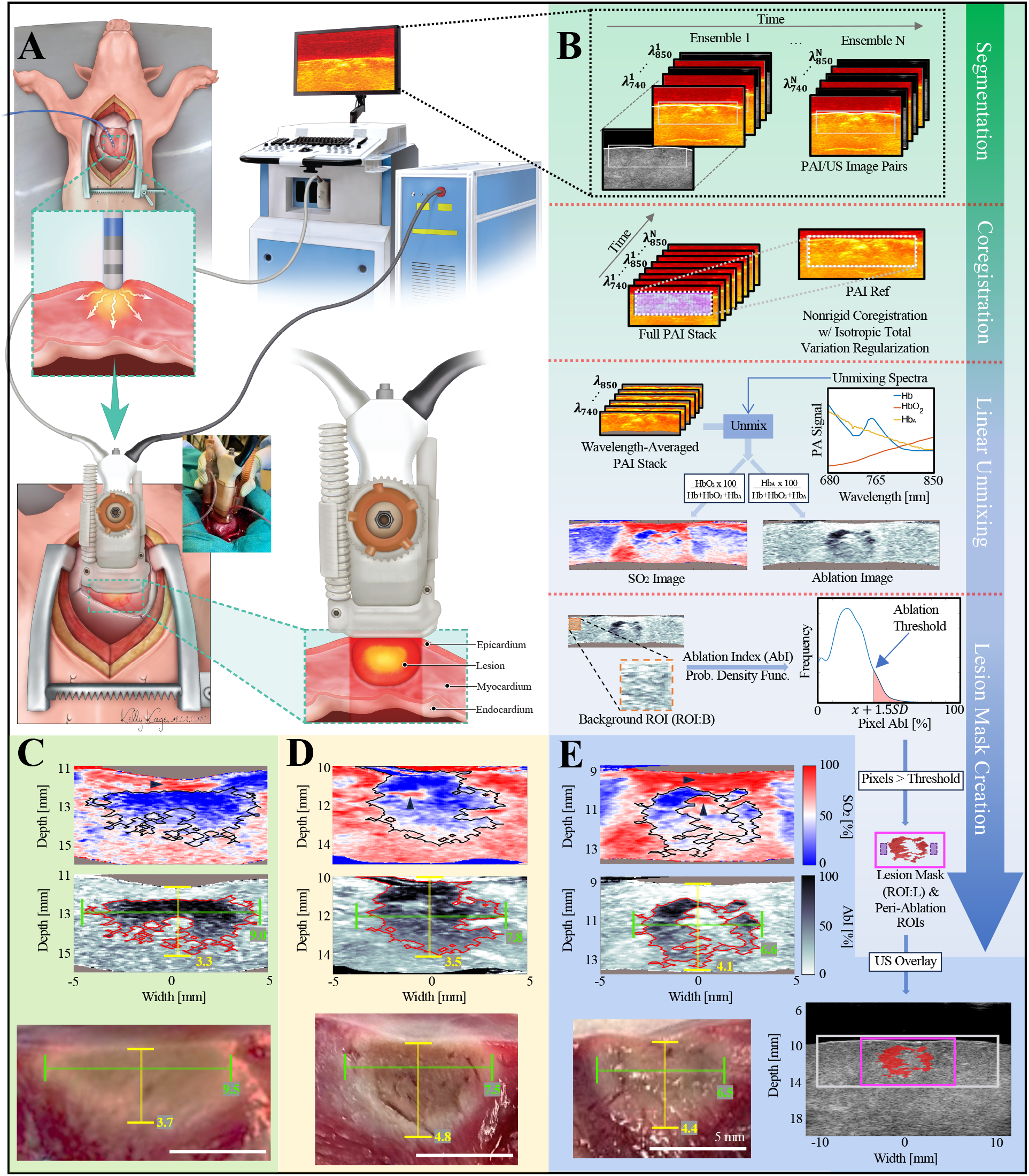
A Open-chest porcine model with RFA over epicardial left ventricle. PAI/US performed on RFA lesion using a vacuum coupler with gelatin stand-off to maintain acoustic coupling and mitigate motion. Experiments were approved by the IACUC at the HMRI (#IS00006601 approved 1/25/22). **B** Post-processing workflow applied to PAI/US data. First, surface *segmentation* conducted on US (white trace) and applied to matched PAI images; 18×5 mm ROI (gray & dashed white boxes) defined for each PAI image. Then, nonrigid *coregistration* (based on PAI reference) applied to full PAI stack. Next, wavelength-averaged PAI stack generated and *unmixed* (HbO_2_, Hb, & Hb_A_) to generate SO_2_ and ablation images. Finally, background ROI (orange box; 2×2 mm) established in upper (i.e., defined by surface segmentation) left-most area of ablation image to calculate *lesion mask creation* threshold; two peri-ablation ROIs (purple boxes; 2×1 mm) defined immediately flanking lesion mask at matched depth center. PAI-based lesion depth based on distance from central tissue surface to maximum mask depth extent, while width based on maximum mask width extent. **C, D**, and **E** show SO_2_ (above), ablation-index (middle), and corresponding stained pathology (below) images of RFA lesions created in 3 porcine subjects. Note all images are displayed with the same scale. Boundaries of ablation depth (yellow) and width (green) are marked in all images with corresponding lesion measurements [mm] provided. **E** (lower right) depicts final post-processing output, where automated thresholding allows for intraoperative US overlaid PAI-based ablation estimate.

An image processing pipeline was developed to mitigate effects of cardiac motion, optimize spectral unmixing, and provide accurate ablation characterization (Fig. 1B). First, the epicardial surface in US/PAI image pairs was segmented from the anechoic stand-off, and a rectilinear region of interest (ROI) defined from the segmented surface for each PAI image. Next, normalized 2D cross-correlation (CC) was calculated using each of these ROIs as a search kernel within the other images of that acquisition. Nonrigid image coregistration was achieved with a minimization approach using isotropic total variation regularization to parametrically register PAI images to a reference image (i.e., highest mean CC)^3^; only images with post-registered CC >0.6 were retained. Following coregistration, PAI images of matched wavelength were averaged, and linear unmixing performed for three chromophores: Hb, HbO_2_, and Hb_A_ (see “Unmixing Spectra” in Fig. 1B; published Hb_A_ spectrum used^2^). Unmixed images representing oxygen saturation (SO_2_=100*HbO_2_/[Hb+HbO_2_+Hb_A_]) and ablation index (AbI=100*Hb_A_/[Hb+HbO_2_+Hb_A_]) were generated for each lesion. Lastly, the ablation-index image was binarized using a threshold 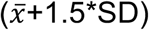 established from an unablated background ROI (ROI:B), creating a lesion mask (ROI:L) for ablation depth and width estimation (Fig. 1C-E) and to establish two peri-ablation ROIs. Separately, while blinded to these results, lesion depth and width were manually determined in stained gross pathology images. Metrics are reported as 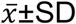, while a paired, two-tailed t-test (α=0.05) was used for comparisons.

Areas of central tissue necrosis were coincident with increases in unmixed Hb_A_. No significant differences were found between PAI-based and gross pathology lesion dimensions (Fig. 1C-E) by depth and width (|Δ| of 0.7±0.5 mm, *p*=0.17, and 0.3±0.2 mm, *p*=0.85, respectively). Average lesion, peri-ablation, and background AbI are 58.8±7.0, 21.4±2.5, and 24.1±3.0%, respectively, and SO_2_ are 32.6±7.8, 61.1±9.1, and 42.6±18.6%, respectively. Further, an average contrast-to-noise ratio 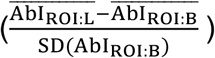 of 2.8±0.2 demonstrates lesion detectability. Although not significant, PAI-based depth tended to underestimate pathology depth, which could be due to transducer compression or elevational transducer tilt (i.e., not perpendicular to surface). During RFA, denaturation of proteins and thermal disruption of vascular membranes cause interstitial edema accompanied by (intracapillary) erythrocyte accumulation and extravasation,^4^ which could account for superficial and focal regions of increased HbO_2_ noted within the ablation regions (triangle markers in Fig. 1C-E).

These preclinical data provide initial proof of concept and feasibility of intraoperative PAI/US guidance of RFA for cardiac arrhythmia, which could allow for more effective, adaptive treatments to reduce recurrence rates. With the addition of a pulsed-laser source, PAI data could be obtained via modified intracardiac ultrasound, allowing translation without marked changes in clinical workflow. Further, catheter-based PAI RFA systems have been demonstrated, advancing PAI’s clinical feasibility.^5^ Future directions include assessment of endocardial lesions, PAI of intraoperative lesion progression, and identification of ablation “gaps” to guide directed RFA and improve clinical procedures.

## Acknowledgments

The authors are grateful to Yareli Carcamo-Bahena, Amber Lee Royal, Houston Methodist Institute for Technology, Innovation & Education (MITIE^SM^), Mr. Daryl Schulz from the Preclinical Catheterization Core, and the Comparative Medicine Program at HMRI.

## Data availability statement

Data that support the findings of this study are available from the corresponding authors upon reasonable request.

